# Impact of the COVID-19 pandemic on mental healthcare consultations among children and adolescents in Norway: a nationwide registry study

**DOI:** 10.1101/2021.10.07.21264549

**Authors:** Miriam Evensen, Rannveig Hart, Anna Aasen Godøy, Lars Johan Hauge, Ingunn Olea Lund, Ann Kristin Skrindo Knudsen, Maja Weemes Grøtting, Pål Surén, Anne Reneflot

**Affiliations:** Department of Health and Inequality, Norwegian Institute of Public Health, Oslo, Norway; Institute for Social Research; Centre for Evaluation of Public Health Measures, Norwegian Institute of Public Health, Oslo, Norway; Department of Health Services Research, Norwegian Institute of Public Health, Oslo, Norway; Department of Mental Health and Suicide, Norwegian Institute of Public Health, Oslo, Norway; Department of Mental Disorders, Norwegian Institute of Public Health, Oslo, Norway; Department of Psychology, University of Oslo, Oslo, Norway; Centre for Disease Burden, Norwegian Institute of Public Health, Oslo, Norway; Department of Child Health and Development, Norwegian Institute of Public Health, Oslo, Norway; Department of Health Management and Health Economics, University of Oslo

## Abstract

The COVID-19 pandemic and its associated restrictions may have affected children and adolescent’s mental health adversely. We cast light on this question using primary and specialist consultations data for the entire population of children 6-19 years in Norway (N=908 272). Our outcomes are the monthly likelihood of having a consultation or hospitalization related to mental health problems and common mental health diagnoses. We compared a pandemic (2019-2021) to a pre-pandemic (2017-2019) cohort using event study and difference-in-difference designs that separate the shock of the pandemic from linear period trends and seasonal variation. We found temporary reductions in all mental health consultations during lockdown in spring 2020. In fall 2020 and winter 2021, consultation volumes in primary care increased, stabilizing at a higher level in 2021. Consultations in specialist care increased from spring 2021. Our findings could suggest a worsening of mental health among adolescents.

## INTRODUCTION

The COVID-19 pandemic, declared by the WHO on March 11, 2020, prompted a range of interventions such as social distancing and stay-at-home orders that affected the everyday routines for children and adolescents, including closing of schools and leisure time activities to slow transmission rates. In Norway, a national eight-week lockdown was implemented from March 12, with gradual re-opening throughout the summer of 2020. However, as the pandemic continued during 2020 and 2021, many social restrictions were sustained and reinforced. There have been widespread concerns amount the impact of these restrictions on children’s mental health (1–3).

Childhood and adolescence are a peak time for the onset of common mental health problems such as anxiety, depression, and ADHD (4). Estimates show that about one in five children and adolescents in Western countries suffer some impairment from mental health problems (5). If left untreated, mental health problems can have lasting effects into adulthood and are associated with lower education and income (6,7).

Several factors could potentially worsen mental health among children and adolescents in the wake of the pandemic. For example, stay-at-home orders, including school-closings and restrictions on leisure activities and social gatherings, could lead to increased loneliness and isolation with potentially harmful consequences (8,9). Uncertainty about the length and scope of the pandemic may also lead to fear and worries (10). The pandemic was followed by an economic downturn, including job loss and economic uncertainty, known to have adverse effects on children’s mental health (11,12). Transitions to homeschooling during the pandemic negatively influenced many children’s learning outcomes which may spill over to their wellbeing (13). Furthermore, as mental health problems are more prevalent among children of lower socioeconomic origins, social distancing measures may exacerbate already marked social inequalities in child health (14). In contrast, there are reports of unintended benefits of the pandemic, such as reduced bullying (15), reduced parental stress (16), and increased awareness about mental wellbeing, which could buffer against some detrimental consequences.

Shortly after the onset of the pandemic, there were international reports of a possible worsening of mental health among children and adolescents (17). However, most of these studies were based on convenience sampling, relied on cross-sectional estimates on measures of mental health, and focused on mental health problems during quarantine (18). Even before the pandemic, rates of mental health problems had been increasing (19,20), urging caution in attributing any increase to the pandemic and its associated restrictions. Existing evidence from larger studies comparing measures of mental health collected before the pandemic with data collected during the pandemic is mixed (10,21,22). Two studies of short-term consequences (up to summer 2020) show no substantial changes in mental health (21,23). Two studies follow children to fall 2020, an Icelandic study report deterioration in children’s mental health while a Norwegian study suggests no substantial changes (24,25). Beyond differences in the observation period, the mixed findings may reflect differences in questionnaire scales, age profiles, sample selections, and settings. Moreover, previous studies have relied on self- or parent-reported symptoms of mental health problems with less knowledge about healthcare use for mental health problems. The latter is important, as changes in healthcare utilization, particularly specialist healthcare, for mental health problems may indicate a more severe change in children and adolescents’ mental health status than can be captured through symptom questionnaires. Finally, the pandemic and its associated restrictions may have had both acute and longer-term impacts on mental healthcare utilization, which may have differed as the pandemic evolved. For example, reduced capacity or fear of contagion may have reduced utilization. In contrast, an increased focus on mental health may have lowered the threshold for seeking professional help. In the acute phase, mental healthcare may have been reduced, for instance, due to lockdown or fear of contamination of the virus. This may in turn, lead to a longer-term increase in mental healthcare utilization as a possible result of a previously unmet need for mental healthcare utilization, an increase in mental health problems, or a reduced threshold for help-seeking.

This study examines changes in consultation volumes related to mental health symptoms and disorders among children 6-19 years old using population-wide data on all primary and specialist healthcare use during the pandemic compared to pre-pandemic years. Our approach allows us to net out seasonal effects and period changes. Further, we examined whether consultation volumes changed more among children with high and low parental SES. In Norway, primary and specialist healthcare is free for all children below 18 years old and mental healthcare for children has been operating at normal capacity.

## DATA AND METHODS

### Data sources and study population

We used data from the Norwegian registry BeredtC19, a national emergency preparedness registry administered by the Norwegian Institute of Public Health (26). It includes data from the Norwegian Control and Reimbursement Database (KUHR) and the National Patient Registry (NPR) matched with data from the Population Registry (Statistics Norway). Unique (de-identified) personal identifiers allow for linkage between different registries and between children and their parents. The study sample was restricted to all children aged 6-19 in 2018 or 2020 (see Appendix A for details on sample construction).

### Health service use for mental health problems

Diagnoses of mental health problems were taken from two sources: reimbursement data from primary healthcare services (KUHR) and specialist data from the NPR. Primary healthcare comprises services such as consultations with general practitioners (GPs) and emergency room visits. Diagnostic information is registered in KUHR according to the International Classification of Primary Care (ICPC-2) with either a symptom or disorder code (27). The NPR is a nationwide registry covering all consultations in specialist healthcare coded in accordance with the 10^th^ edition of the International Classification of Diseases and Related Health Problems (ICD-10). A referral from the GP is necessary to get specialist treatment (except for acute hospitalizations).

Monthly measures indicating at least one mental health consultation or hospital admission were constructed for: (i) all mental symptoms and disorders registered in primary care and specific diagnoses for ADHD, anxiety, depression, and sleep problems. (ii) all mental disorders in specialist care as well as specific diagnoses for ADHD, anxiety, depression, and hospitalizations (see Table 1 for details on coding). Due to high levels of comorbidity between anxiety and depression, we analyzed these disorders jointly (28).

**Table 1:**
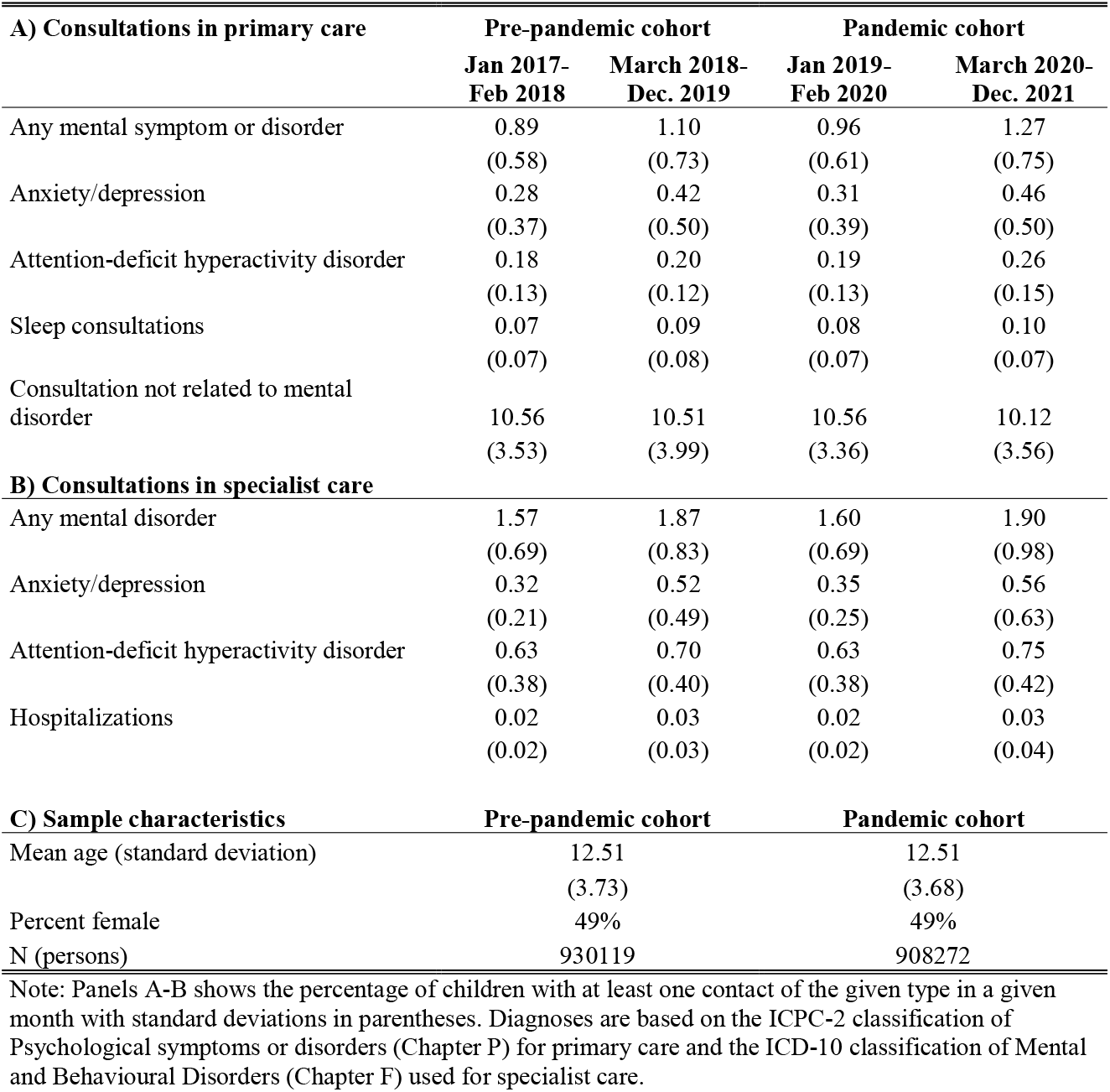
Descriptive statistics on consultations for mental health symptoms and disorders in primary and specialist healthcare and individual characteristics for Norwegian children 6-19 years old.

### Covariates

All models control for (or are estimated separately by) sex (female=1, male=0) and age category (ages 6–12, 13–15, and 16–19), which corresponds to the age of enrollment in primary, secondary, and high school. We also report analyses stratified by the capital area (counties Oslo and Viken) versus all other geographic regions in Norway. The geographic regions are identified using municipality identification numbers. Information on these variables was taken from the Population Registry. We also report analyses stratified by parental occupation as a measure of social background. Parental occupational codes are taken from the Employer-Employee Registry, and were measured using the International Standard Classification of Occupations, ISCO-88 (21). Using the information on the parent with the lowest first digit in the ISCO code, corresponding approximately to the highest occupational status, we distinguish between three main parental class categories: upper white-collar, lower white collar and blue-collar.

### Statistical methods

To evaluate how consultation volumes in primary and specialist healthcare for mental symptoms and disorders changed due to the COVID-19 pandemic and its countermeasures, we compare the use of health services from January 2019 to December 2021 for a pandemic cohort and from January 2017 to December 2019 for a pre-pandemic cohort.

To separate the effects of the pandemic from other temporal trends, we followed the pre-pandemic cohort over the same period and ages, albeit two calendar years earlier. We show bivariate trends for the pre-pandemic and the pandemic cohorts. We fit multivariate event study models with controls for month and time in years to formally test, month by month, whether the use of healthcare services in the period 2019-2021 differs from two years earlier. Data for the first part of the period is used to assess whether trends were comparable in the two cohort groups before the onset of the pandemic. Data up to February 2020 for the pandemic sample, and February 2018 for the pre-pandemic sample, are used for this purpose. Then, we assess whether diverging trends in consultations emerged at the onset of and during the pandemic and its associated restrictions. To quantify the magnitude of the effects, we also estimate difference-in-difference models, where we group the months into four periods (methodological details in Appendix A). An increase in consultations could be due to more children having consultations, longer treatment durations, or both. To distinguish these two drivers, we also calculated simple difference-in-difference models of the change in the number of children with any consultations in the pandemic cohort, relative to the pre-pandemic cohort.

## RESULTS

### Descriptive results

Table 1 shows the distribution of our covariates for a data set of person-months for the entire study. In the pandemic cohort in the period before lockdown (column January 2019-February 2020), 0.96 percent of the children had a mental health consultation in primary care in any given month (Panel A), and 1.6 percent had a mental health consultation in specialist care (Panel B). Females constitute 49% and the mean age at the start of the observation period is 12.51, both in the pre-pandemic and pandemic cohort sample (Panel C).

Figure 1 shows the monthly percentages with at least one consultation for mental disorders from Jan 2019 to December 2021 in the pandemic cohort (full lines), compared to the similar percentages in the pre-pandemic cohort for January 2017 to December 2019 in each age group (dashed lines).

**Figure 1:**
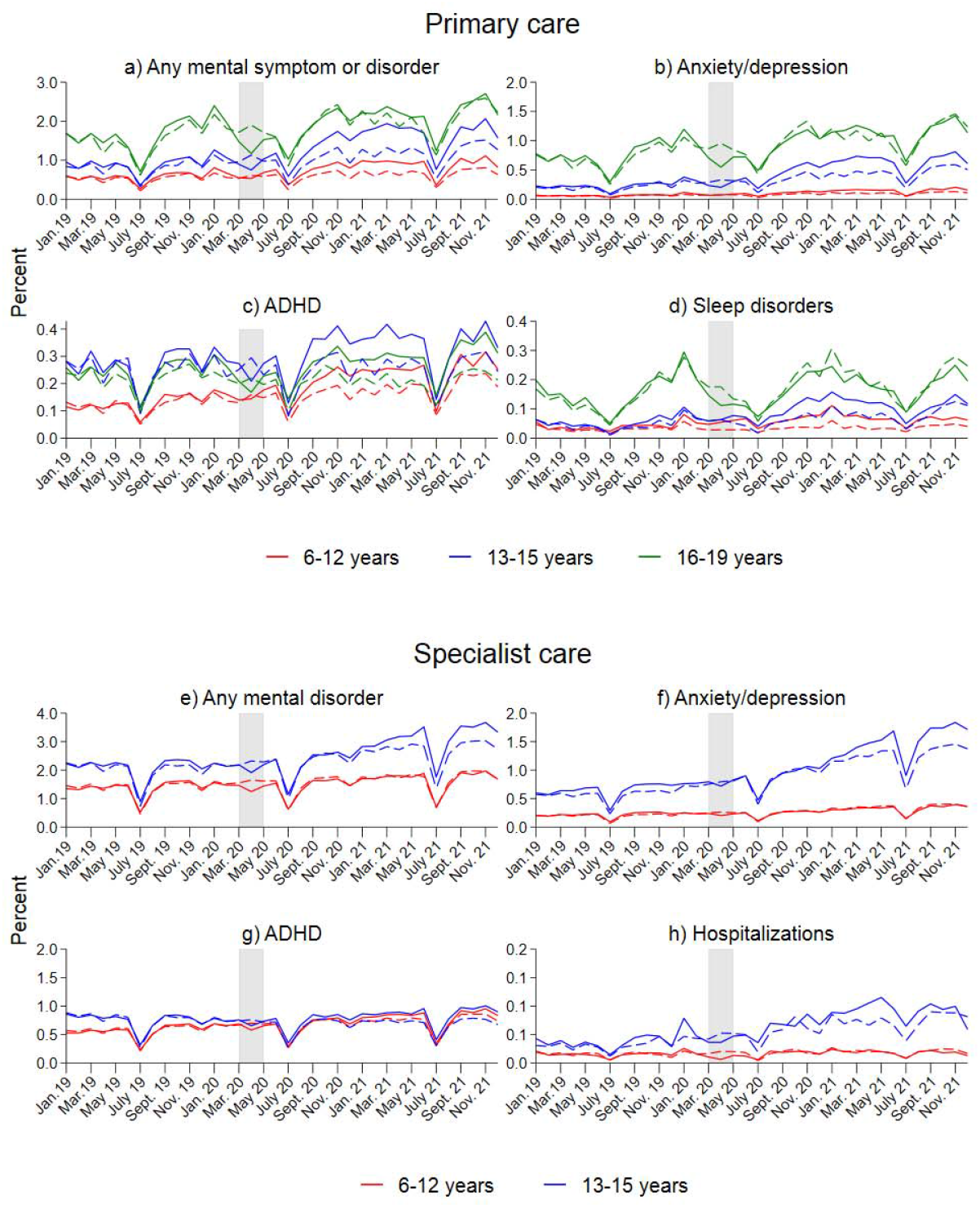
Percent of children with at least one consultation for mental health problems/disorders in primary and specialist healthcare in a given month. Diagnoses are based on ICPC-2 codes for primary care and ICD-10 codes for specialist care (see Table A.1 for the full list of diagnostic codes). Separate calculations by age and treatment group. The shaded area indicates the full lockdown period. The x-axis refers to the measurement time for the main sample (full lines). Dashed lines refer to the comparison groups, observed from January 2017-March 2019. For the comparison sample, all measurements are made 24 months earlier.

First, the graphs document marked seasonal variations in consultations, with large decreases in July each year (school holiday) and small peaks in January for some outcomes. Up to March 2020 (March 2018), the trends are comparable in the pandemic and pre-pandemic cohorts. Second, there is a weak increase in consultations over time, so the pandemic cohort is often at a higher level than the pre-pandemic cohort until March 2020. Third, the percentage of consultations dropped sharply around the lockdown in the pandemic cohort but increased rather quickly to pre-lockdown levels. Finally, from September 2020, the percentage of consultations started to increase faster in the pandemic cohort compared to the pre-pandemic cohort. The increase pertains to all primary care outcomes (Panels a-d). For specialist care (panels e-h), there is a tendency of a faster increase from January 2021 for the age group 13-15 years, persisting throughout 2021.

### Multivariate results

To formally test whether the healthcare utilization of the pandemic cohort differed from that of the pre-pandemic cohort, we estimated event study models, netting out shared seasonal differences and secular change over time.

Figure 2, panel a, shows that the monthly probability of having any primary healthcare visits related to mental health decreased at the start of lockdown. The dip is largest for anxiety and depression consultations (Panel b). After the lockdown period, we see a quick rebound and leveling off, with a slight increase in summer relative to the pre-pandemic cohort. As indicated by the bivariate plots (Fig. 1), the percent of children with a primary care consultation (Fig. 2, Panel a), as well as for all diagnostic groups in primary care (Panels b-d) starts to increase faster around August 2020 for the two youngest age groups, levelling off at a substantially higher level at the end of 2021. There is a significant increase among 13-15-year-old children for specialist consultations for ADHD (Panel g) and hospitalizations (Panel h) in 2021. We see the same tendency for specialist consultations for any mental disorder (Panel e) and anxiety/depression (Panel f), albeit these are less precisely estimated

**Figure 2:**
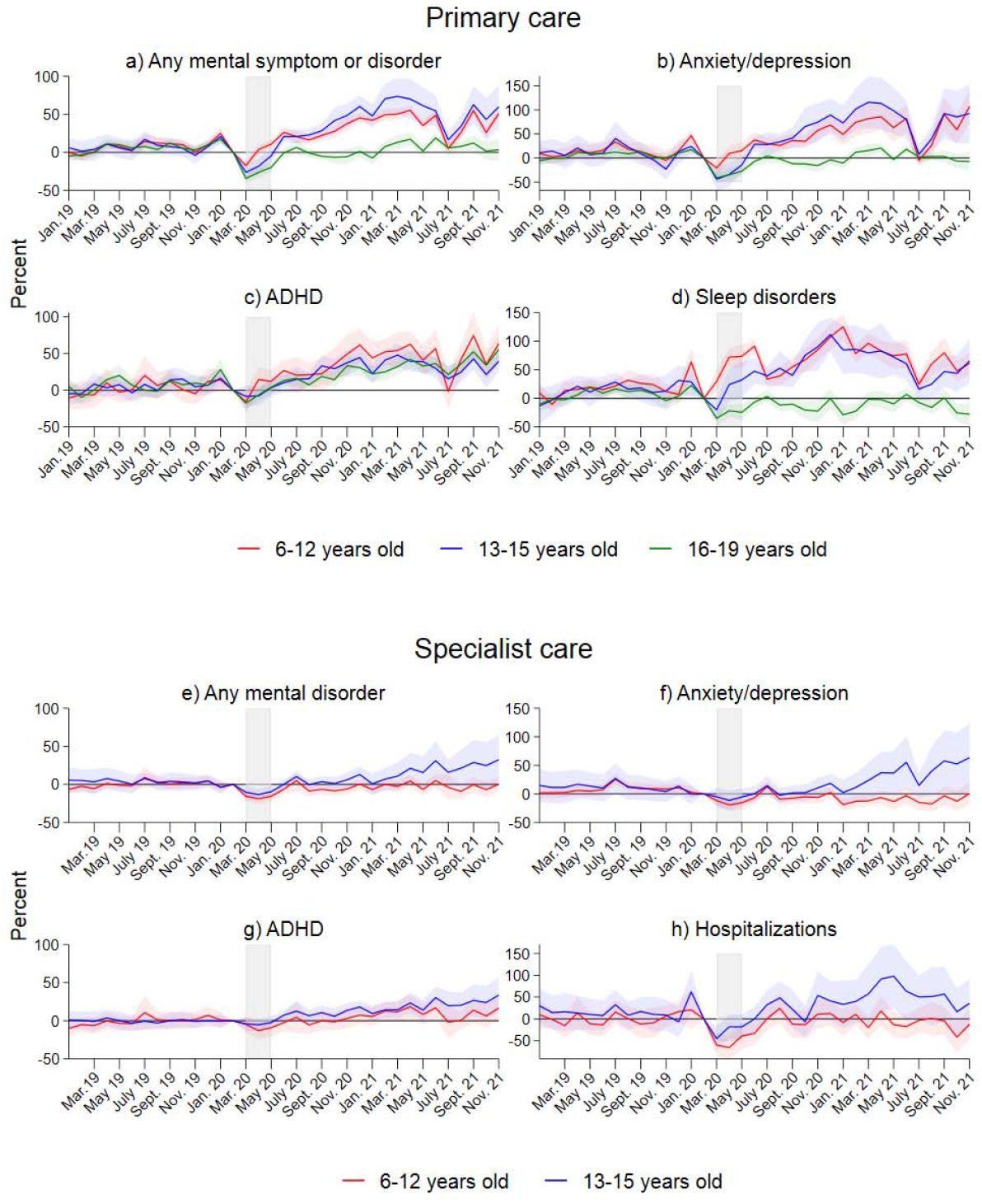
Results from separate event study models for three age groups. Complete lines show coefficients and shaded areas their 95% confidence intervals. Coefficients and confidence intervals are scaled to the pre-lockdown level in the main sample (see Table 1). The outcome is the monthly propensity to have at least one consultation of the type mentioned in the panel headers. Diagnoses are based on ICPC-2 codes for primary care and ICD-10 for specialist care (see Table A.1). The x-axis refers to the measurement time for the main sample. For the comparison sample, all measurements are taken 24 months earlier. Models control for the duration in years, sex, municipality, month, and Easter holidays.

To understand the magnitude of these effects, we show difference-in difference estimates (ie., average monthly coefficients) across 4 time periods (lockdown, summer 2020, fall 2020, and the year 2021) in Table 2.

**Table 2:**
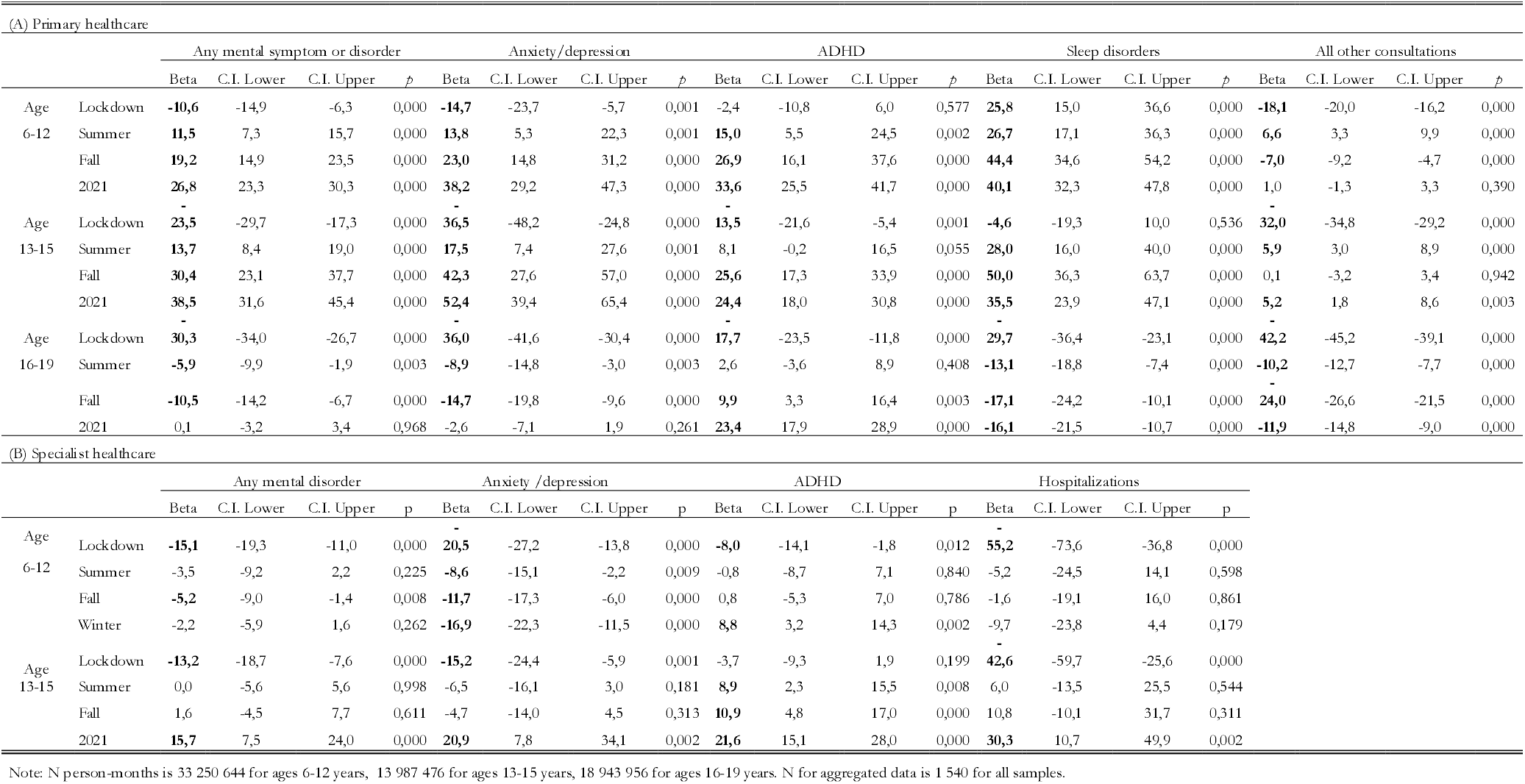
Difference-in-difference estimates of change in the monthly probability of healthcare consultations with 95% CI.

The estimates for all primary mental health consultations during lockdown suggest a 30.3 percent reduction among 16-19-year-olds, 23.5 percent reduction for those aged 13-15, and 10.6 percent reduction for those aged 6-12, all statistically significant at the 5 percent level. During 2021, however, the percentage of children aged 13-15 years with any mental health consultation is 38.5. percent higher in the pandemic cohort than the pre-pandemic cohort. With a baseline of 1.1 percent before the pandemic, this increases to 1.5 percent with a monthly consultation. For 6–12-year-old children there was an increase of 26.8 percent for all mental health consultations. With a baseline of 0.7 percent before the onset of the pandemic, this gave a level of 0.9 percent after the onset of the pandemic. Except for ADHD consultations, there is no increase for the oldest age group.

For 13-15-year-old children, the percentage with any specialist consultation increased with 15.7 percent in 2021 compared to pre-pandemic levels. There was no statistically significant increase for 6-12 year old children. The increase in hospitalizations among 13-15-year-olds is estimated to be 30.3 percent in the difference-in-difference models. ADHD consultations in specialist care increased in both age groups. Among 6-12-year-old children, an increase of 8.8 percent from a 0.6 baseline gave a level of 0.7 percent after the onset of the pandemic. For children aged 13-15 years, the increase was 21.6 percent. Combined with a baseline of 0.7 percent, this gave a level of 0.8 percent after the onset. Specialist consultations for anxiety and depression increased with 20.9 percent among 13-15-year-old children, and fell with 16.9 percent among 6-12 year old children.

Finally, we show simple difference-in-difference estimates for the change in number of children in contact with health services. For primary care, the estimates in Table A.2 are comparable to the main results. For specialist care, estimates for the number of children are consistently lower than the main results, suggesting that longer treatment for those already in care is an important driver of the main results.

### Subsample analysis

We ran the models separately by sex (Appendix Figure A.1). Across outcomes in primary care, the increase starting around August 2020 is stronger among females than males, but confidence intervals overlap. The increase in specialist care consultations (Panels e-h) is found among females only. We also estimated the sex-specific models for 13-15-year-old children only, the age group where we have seen the largest effects (Appendix Figure A.2). For this group, the sex differences in increases are even larger, 95% confidence interval for the event study estimates is no longer overlapping for most outcomes, except for sleep disorders (Panel d).

We also split the sample by parent’s socioeconomic status using the information on parental occupation from Statistics Norway. Results were similar across socioeconomic groups (Figure A.3).

Finally, we tested whether the effects differed between the Capital area (regions Oslo and Viken), which had the strictest restrictions, and the rest of Norway (Appendix Figure A.4). We find no evidence that the effects are restricted to the Capital area.

## DISCUSSION

Using population-wide data on mental healthcare for the first 21 months of the pandemic, we found a pronounced increase in primary care consultation volumes related to mental health symptoms and disorders among children that depart from previously established increases over recent years. Although the number of consultations for mental health declined sharply during the initial lockdown period, consultation volumes returned to pre-pandemic levels by June 2020. However, our models uncovered a gradual increase in the number of primary consultations related to mental health during fall 2020 and continuing through 2021, corresponding to the second and third waves of infections and their associated social distancing mandates. In the more selective specialist healthcare, effects manifested somewhat later.

Compared to pre-pandemic years, primary care mental health consultations increased by 38.5 percent in 2021 for 13-15-year-old children. Both on an absolute and relative scale, the increase was highest for anxiety and depression, with a 52.4 percent increase in 2021. ADHD and sleep disorders increased by 24.4 and 35.5 percent, respectively, for the same period and age groups. The increase was found for both sexes but was most pronounced among girls.

The increase in primary care consultations was less pronounced for older adolescents (16-19 years old). It could be that the oldest adolescents are better at coping with the pandemic and its associated restrictions. This would be in line with some previous Norwegian studies but slightly at odds with other studies that have reported the largest mental health deterioration among older children (24,29). A likely cause of the lack of increase, however, is a policy change regarding absences for upper secondary school students. Before the pandemic, absences from upper secondary to illness or injury were required doctor’s certification. This requirement was lifted shortly after the beginning of the pandemic, reducing the demand for primary care consultations in this age group (30).

In Norway, less severe mental health problems will generally be treated by a physician in primary care, while more severe cases will be referred to specialist treatment by the physician and treated by psychologists and psychiatrists. The lag in effects in specialist treatment, compared to primary care, could reflect constraints in treatment. Already before the pandemic, specialist healthcare had long wait lines and delayed access to treatment (31), meaning that increased demand for specialist healthcare would not necessarily show up immediately in our data as an increase in consultations. Among specialist care treatments, psychiatric hospitalizations are not affected by capacity constraints in the same way. For this outcome, we found an increase already from fall 2020.

There have been concerns that the pandemic might increase the already large social inequalities in the prevalence of mental health disorders between children from high and low-income families (14). However, we found that the increase in mental health consultations was largely similar among children of parents with high and low occupations. This may suggest that the pandemic has not exacerbated social inequality related to mental health consultations. It could also be that some families from disadvantaged backgrounds experienced poorer health but could not seek help for various reasons related to the pandemic. If so, there might be concealed differences related to social inequality. For example, we know that unemployment rates became higher among low educated individuals and if these parents experience worse health, they might become less capable of attending to their own children’s needs.

To our knowledge, ours is the first study to examine possible increases in consultation volumes related to children’s mental health with a research design that handles both age change and (linear) period trends. We established empirically that the pre-pandemic and the pandemic cohorts had similar trends in consultation volumes before March 2020 and then showed that the trends diverge markedly over time. Thus, our results suggest that the pattern of consultation volumes increased beyond what we would expect based on previous trends. The increase in primary consultation volumes became visible 6 to 8 months into the pandemic and about a year into the pandemic for specialist consultations. This suggests that most children (and parents) coped with changes in the short run. Still, in the long run, consultation volumes increased.

To the extent that our findings reflect a worsening of underlying mental health, this in line with an Icelandic study indicating that self-reported depression increased during the pandemic (24). In contrast, Norwegian survey-based studies indicate no increase up to fall 2020 (25). However, the conflicting findings may reflect the timing of data collection since our study has a considerably longer follow-up (until the end of 2021), and the largest effects are found toward the end of the period. The decline in consultations during the initial lockdown happened in a period where surveys suggest no worsening of mental health. This suggests that our estimates are driven by a temporary change in healthcare utilization rather than an improvement in underlying health.

There are multiple aspects of the pandemic that could plausibly lead to deteriorating mental health, including diminishing social support networks (32) and unpredictability and disruption in daily routines (33). Even as restrictions eased, public health measures such as social distancing and attempts to reduce the mixing of students across cohorts severely limited social gatherings (34). However, the increase seen in consultations could also be due to greater media coverage and awareness of mental health problems. This could lead parents or physicians to rate children to have more symptoms now compared to in pre-pandemic years. Increased family time during the pandemic (16) could make parents more responsive to their children’s symptoms. If fears of contracting COVID-19 or overburdening the healthcare system increased the threshold for seeking help, the worsening of mental health would be larger than the increase in consultations suggest. Changes in consultation practice, such as more online consultations, could also affect our results by lowering the threshold to contact physicians. However, findings from a sensitivity analysis examining consultations volumes for overall healthcare did not support this (cf. Appendix Fig.2).

There are strengths and limitations to our study. Unlike prior studies that use self-report data, we likely capture clinically relevant symptoms and conditions causing distress in everyday life. Our inclusion of hospitalizations also means that we can examine changes among vulnerable children, which are likely not included in survey-based studies. However, relying on healthcare data also means that we only examine a small proportion of children with mental disorders. To the extent that one is interested in underlying mental health, it is a limitation of our study that our results can also be influenced by other changes affecting health service use, as discussed above. Future studies should examine underlying prevalence trends, and using direct mental health measures in an equally robust design would be particularly valuable.

As for the validity of diagnoses in primary care, a previous study compared interview-based diagnoses for depression and anxiety with diagnoses taken from KUHR and NPR and found that registry-based diagnoses have moderate sensitivity and excellent specificity, with very few false positives (35). While we are not able to rule out that physician evaluations may have changed during the pandemic, we consider it unlikely that the increase is caused by sudden changes in diagnostic practice.

Finally, our results are found in a context with relatively low COVID-19 mortality rates and fewer social restrictions than in other European countries such as England and Germany. In Norway, keeping schools open has been a priority, and an extensive welfare state ameliorates the consequences of the economic downturn (36). Thus, one might expect a larger deterioration in children’s mental health in contexts where social restrictions have been more profound.

### Conclusion

We found that consultations related to mental health symptoms and disorders in primary care increased during fall 2020 and in 2021 over and above increases that occurred in recent years before the pandemic. The corresponding increase in consultations in specialist healthcare happened later, in mid-2021, except for an earlier onset for hospitalizations. It is paramount to understand the underlying drivers of the increase in consultations, and how any worsening of child and adolescent mental health can be mitigated.

## Data Availability

For confidentiality reasons data cannot be shared.

## Declarations

### Availability of data and material

For confidentiality reasons data cannot be shared.

### Code availability

not applicable

### Ethics committee approval

The study has been approved by the Norwegian Regional Committees for Medical and Health Research Ethics (REC), approval number 2021/267200. We confirm that all administrative permissions have been granted to access and use the data for this study.

