## Supplementary data for "Impact of the COVID-19 pandemic on mental healthcare consultations among children and adolescents in Norway: a nationwide registry study"

**Appendix A:**

*Details on methods and sample construction*

Our main sample is all children that turned 6-19 years in 2020, observed from January 2019 to December 2021. Our comparison sample is all children of the same age (turned 6-19 years in 2018), observed 24 months earlier, from January 2017 to December 2019.^[1]^

To formally compare the trend development in the intervention and comparison group, we estimate event study models, taking the following form:

$y_{i,t}={\sum_{k=-14, k!=-1}^{21} X_{Intervention}*1(t-t{0=k)\beta}_{k}}+{\sum_{Y=-1}^{1} \beta_{Year}X_{Year,i, t}}+{\sum_{W=1}^{12} \beta_{Month}X_{Month,i, t}}+ \boldsymbol{\beta}\mathbf{X}$+$\varepsilon$

Where t0 refers to the first month of lockdown, and k is month number. The expression

$X_{Intervention}*$1(t−t0=k) constructs a dummy variable that takes 1 if the observation is in the main sample, and the month is k months away from March 2020, otherwise 0. The omitted reference category, in which all observations in the comparison sample are included, is the month before lockdown. This comparison allows us to net out overall level differences between the main and comparison group, for instance due to increased use of health services across periods. The parameters of interest are βk ’s, which give us month-by-month estimates of how the trend in the main sample deviates from the trend in the comparison group, and relative to the month before lockdown. The parameter estimates for month net out monthly variations shared across cohort and year. To net out period change also within cohorts, we control for calendar year minus the year of time zero, so that -1 denotes 2017 (2019) in the main (comparison) sample, counting up to 1 for 2019 (2021).  Finally, we include a vector of controls **X**, that includes region, dummies for age category (unless models are separate by age category), a dummy for being male (unless models are separate by sex) and a variable running from 0 to 1 showing the proportion of Easter falling into the given month in the given year.

If pre-trends are parallel, i.e., the βk coefficients for before lockdown (negative t’s) should be insignificant and close to zero. Effects of (prolonged) lockdown should then become emergent no earlier than t0. Note that the direction of effects and their drivers may vary over time: while the access to health services was restricted in the immediate lockdown, they were generally accessible in the prolonged period of social distancing that followed.

We also estimate difference-in-difference models (i.e., average monthly coefficients) for the same outcomes, including the same control variables. In these models, we collapse the duration variables into periods. We group the months into four periods (with measurements in the comparison sample always taken 24 months earlier): lockdown (March-May 2020), summer (June-August 2020), fall (September-December 2020) and 2021 (January-December 2021).

Time varying covariates (parental occupation and age) were measured January 1, 2020, for the pandemic cohort, and January 1, 2018, for the pre-pandemic cohort.

**Appendix B: Supplementary results**

| Table A1: Codes and percent children with mental health diagnoses in primary and specialist care | | | |
| --- | --- | --- | --- |
| **Primary care** | **2017** | **2019** | **ICPC-2 code** |
| Any mental symptom or disorder | 6.45 | 6.93 | All Chapter P codes |
|  | (2.98) | (3.18) |  |
| Anxiety/depression consultations | 2.03 | 2.24 | P74, P76, P79, P82, |
|  | (2.29) | (2.44) | P01, P02, P03 |
| Attention-deficit hyperactivity disorder | 1.28 | 1.35 | P81 |
|  | (0.47) | (0.54) |  |
| Sleep consultations | 0.58 | 0.64 | P06 |
|  | (0.48) | (0.49) |  |
| All consultations | 61.33 | 61.73 | All remaining codes |
|  | (5.91) | (6.57) |  |
| **Specialist care** |  |  | **ICD-10 Code** |
| Any mental disorder | 4.41 | 4.83 | All Chapter F codes |
|  | (1.02) | (1.30) |  |
| Anxiety/depression consultations | 0.96 | 1.13 | F32, F33, F40, F41, F43, |
|  | (0.84) | (0.97) | F93.0, F93.1, F93.2 |
| Attention-deficit hyperactivity disorder | 1.51 | 1.62 | F90 |
|  | (0.32) | (0.37) |  |
| Hospitalizations | 0.20 | 0.24 | All inpatient codes related to mental disorders |
|  | (0.11) | (0.19) |  |
| Note: The table gives the percentage of children that had at least one contact of the type in the given year. Diagnoses are based on the ICPC-2 classification on Psychological symptoms or disorders (Chapter P) for primary care and the ICD-10 classification of Mental and Behavioural Disorders (Chapter F) used for specialist care. | | | |

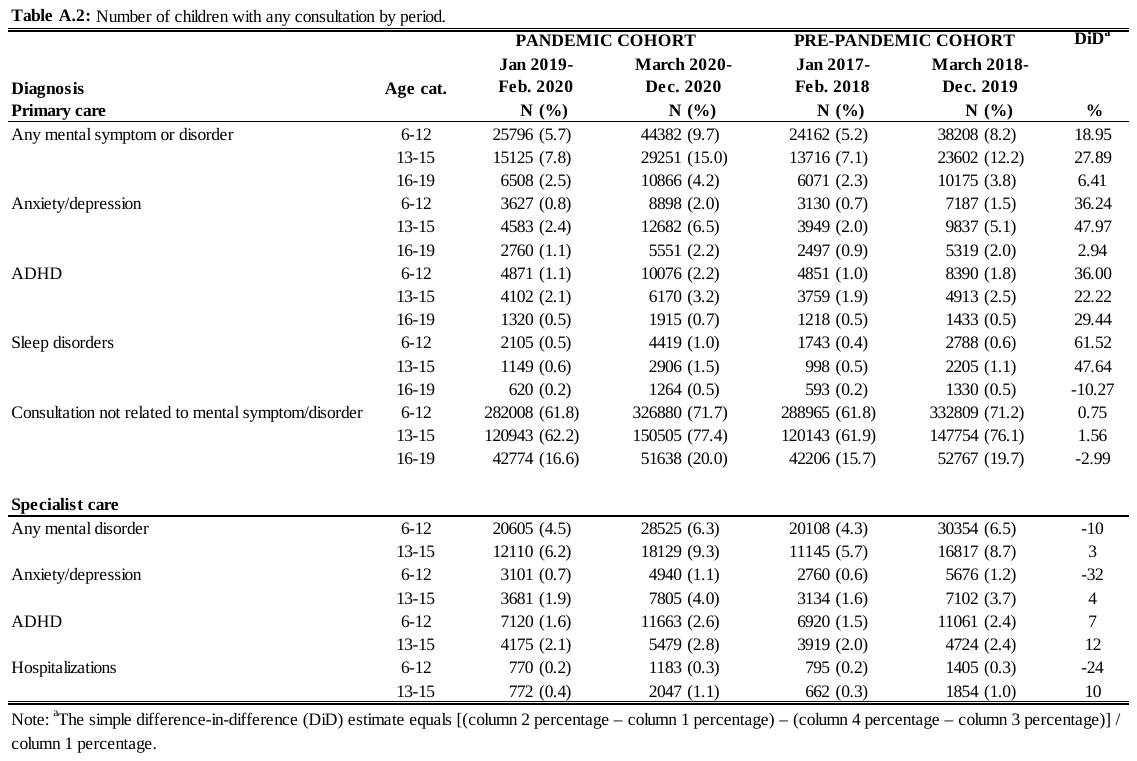

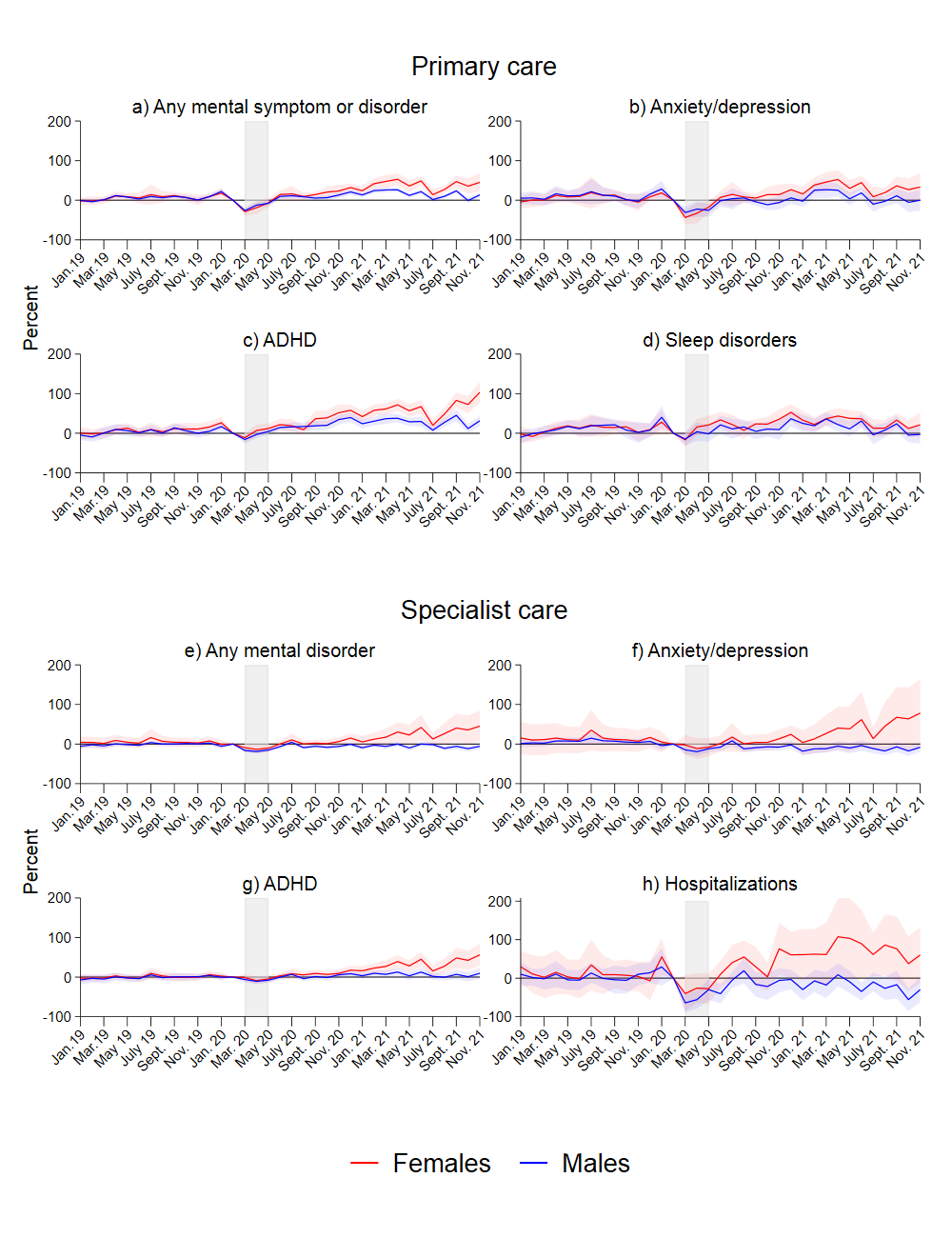
**Figure A.1:** Results from separate event study models for males and females. Complete lines show coefficients, and shaded areas their 95% confidence intervals. Coefficients and confidence intervals are scaled to the pre-lockdown level in the main sample (see Table 1). The outcome is the monthly propensity to have at least one consultation of the type mentioned in the panel headers. Diagnoses are based on ICPC-2 codes for primary care, and ICD-10 for specialist care (see Table A.1). The x-axis refers to the measurement time for the main sample. For the comparison sample, all measurements are taken 24 months earlier. Models control for duration in years, age category, municipality, month and easter.

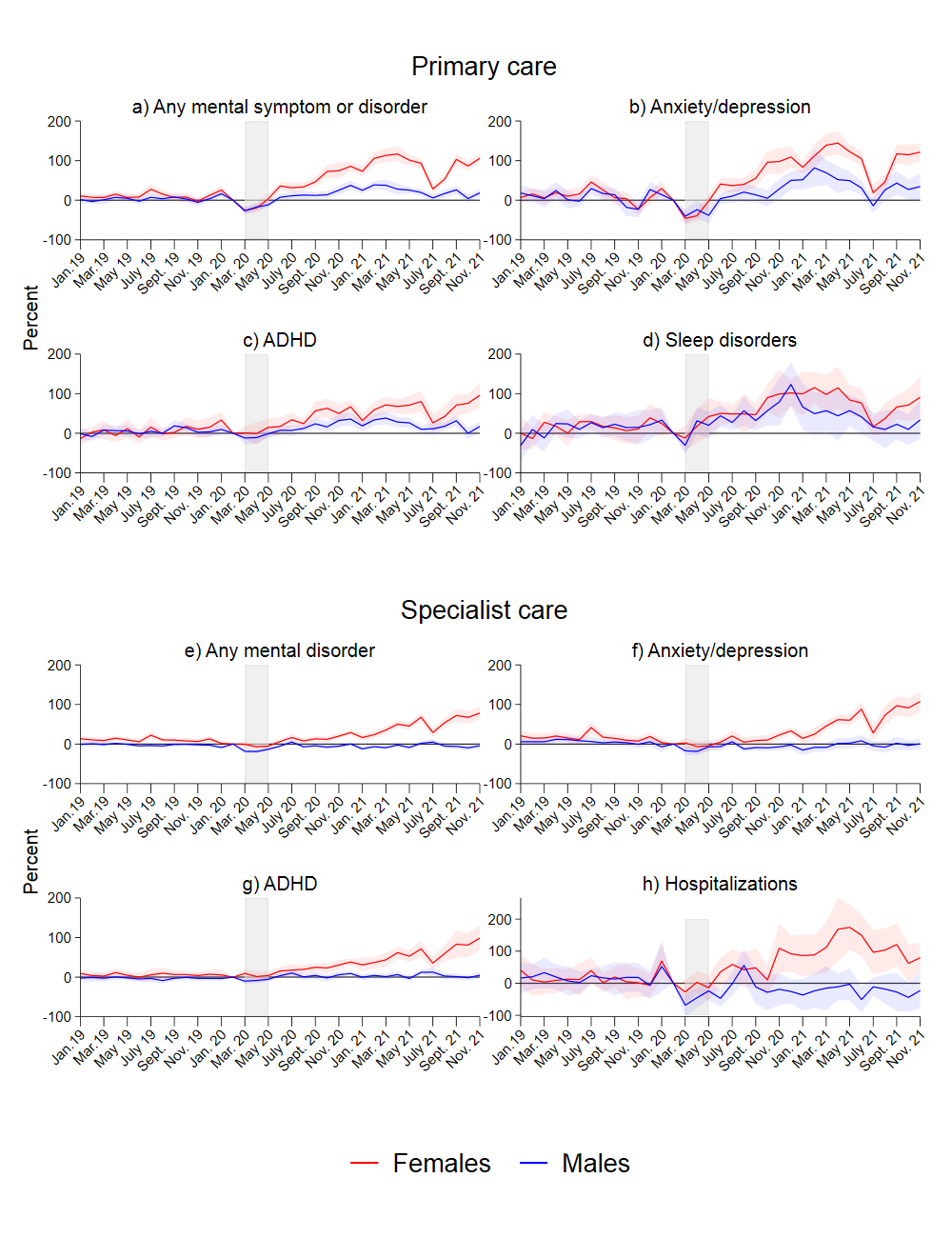
**Figure A.2:** Results from separate event study models for males and female, ages 13-15. Complete lines show coefficients, and shaded areas their 95% confidence intervals. Coefficients and confidence intervals are scaled to the pre-lockdown level in the main sample (see Table 1). The outcome is the monthly propensity to have at least one consultation of the type mentioned in the panel headers. Diagnoses are based on ICPC-2 codes for primary care, and ICD-10 for specialist care (see Table A.1). The x-axis refers to the measurement time for the main sample. For the comparison sample, all measurements are taken 24 months earlier. Models control for duration in years, age category, municipality, month and easter.

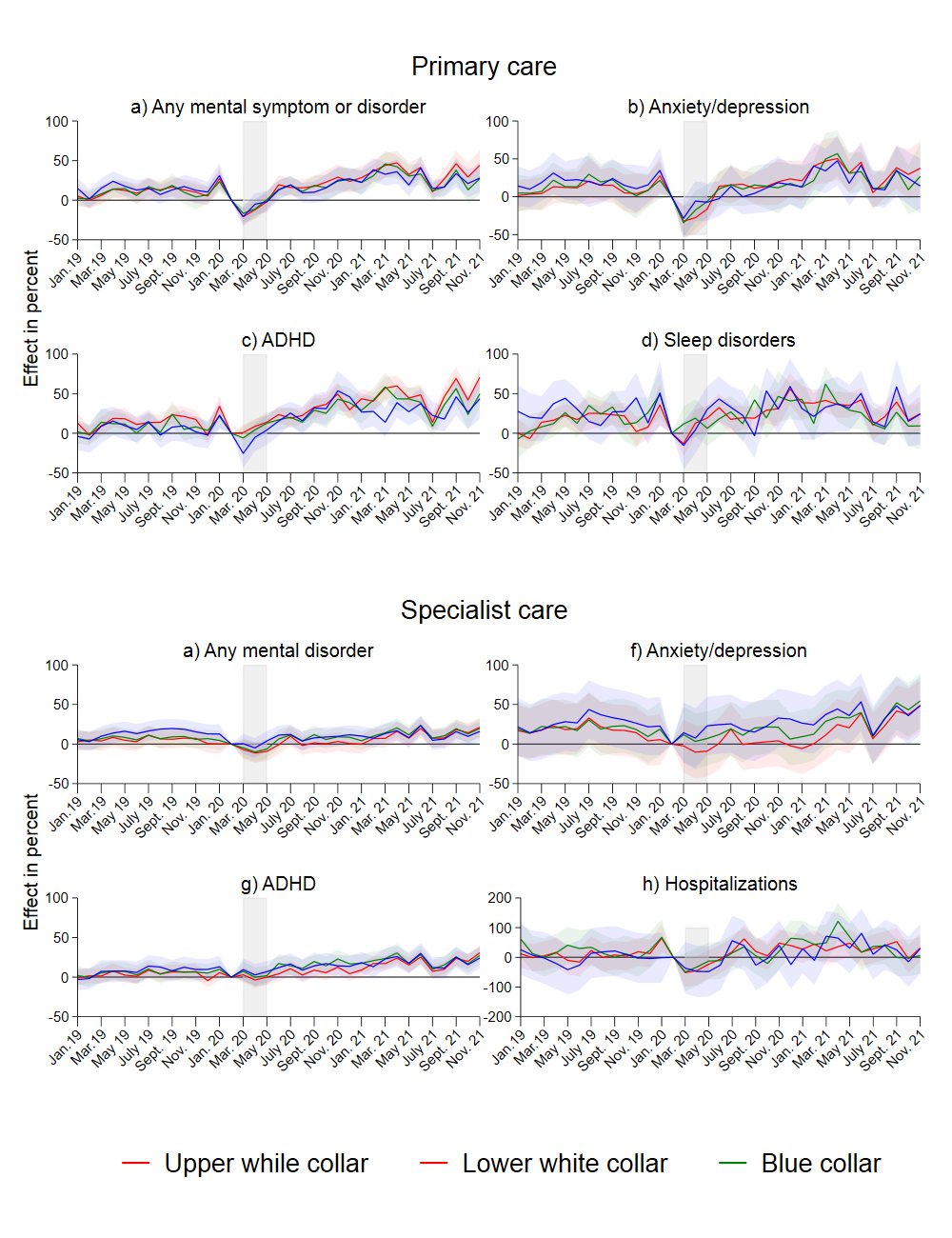
**Figure A.3:** Results from separate event study models by parents’ social background. Complete lines show coefficients, and shaded areas their 95% confidence intervals. Coefficients and confidence intervals are scaled to the pre-lockdown level in the main sample (see Table 1). The outcome is the monthly propensity to have at least one consultation of the type mentioned in the panel headers. Diagnoses are based on ICPC-2 codes for primary care, and ICD-10 for specialist care (see Table A.1). The x-axis refers to the measurement time for the main sample. For the comparison sample, all measurements are taken 24 months earlier. Models control for duration in years, sex, municipality, month, age category and easter.

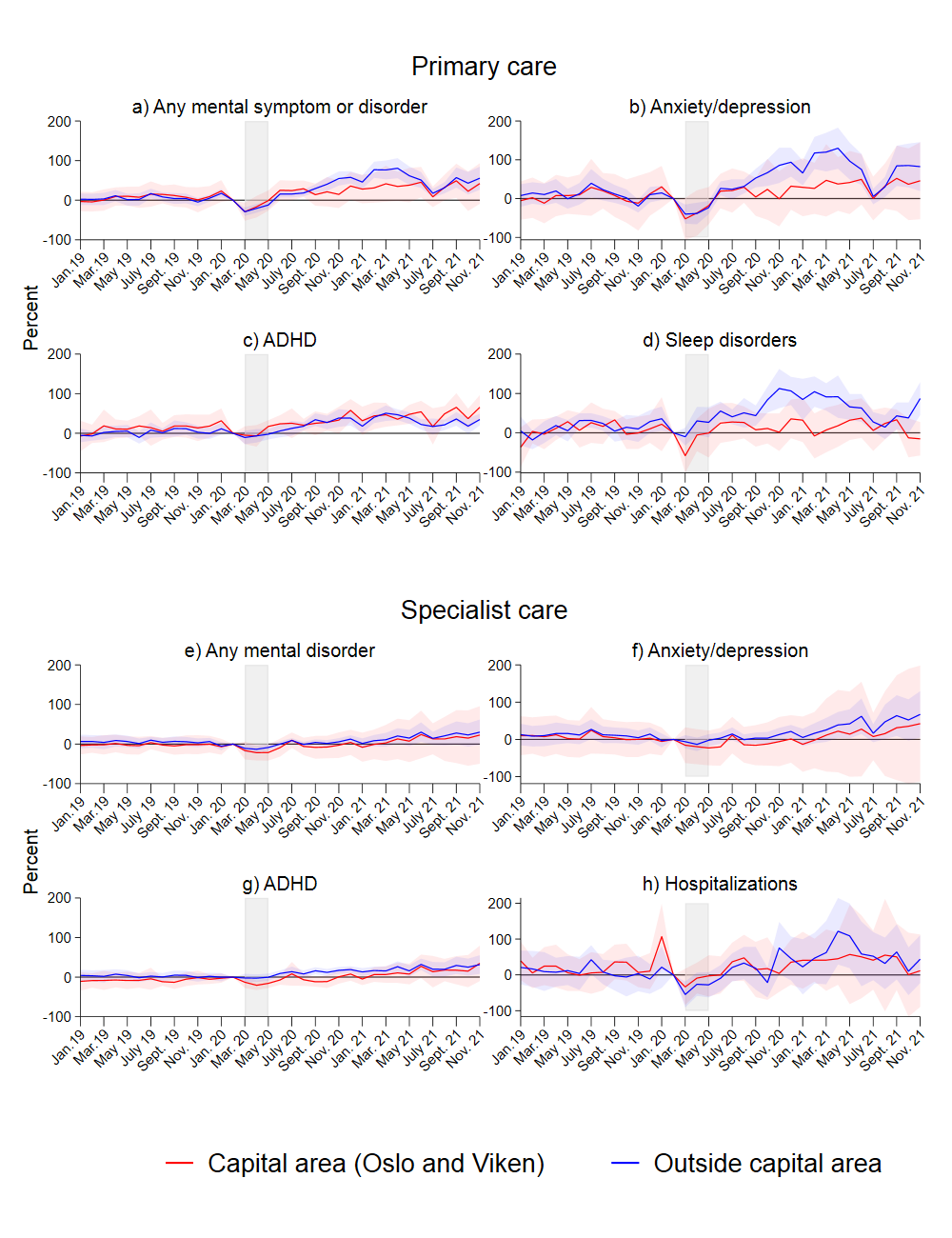

**Figure A.4:** Results from separate event study models for capital area (Oslo and Viken counties) and the rest of Norway, all age groups. Complete lines show coefficients, and shaded areas their 95% confidence intervals. Coefficients and confidence intervals are scaled to the pre-lockdown level in the main sample (see Table 1). The outcome is the monthly propensity to have at least one consultation of the type mentioned in the panel headers. Diagnoses are based on ICPC-2for primary care, and ICD-10 for specialist care (see Table A.1). The x-axis refers to the measurement time for the main sample. For the comparison sample, all measurements are taken 24 months earlier. Models control for duration in years, sex, age category, municipality, month and easter.

*Sensitivity tests*

A concern is that the pandemic and the associated consequences changed all primary health care utilization, so all health care utilization increased, not only that related to mental health. To provide a robustness check of the results for primary health care service use, we show the development for all primary care consultations in Appendix Figure A.5.

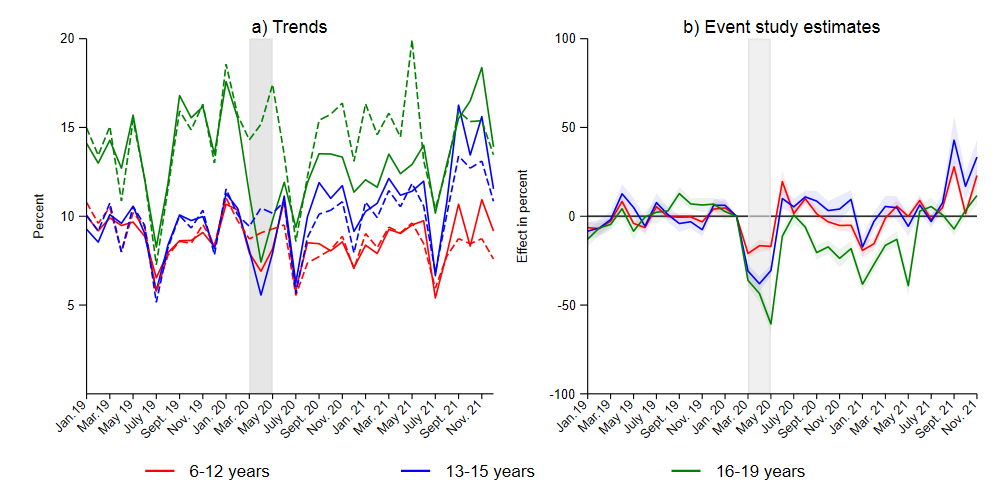

**Figure A.5:** Trends and event study models for any primary care consultation. In Panel b, complete lines show coefficients, and shaded areas their 95% confidence intervals. Coefficients and confidence intervals are scaled to the pre-lockdown level in the main sample (see Table 1). Diagnoses are based on IPCD codes for primary care, and. ICD-10 for specialist care. The x-axis refers to the measurement time for the main sample. For the comparison sample, all measurements are taken 24 months earlier. Separate models by three age groups, primary school (ages 6-12), secondary (ages 13-15) and high school (ages 17-19). Models control for duration in years, sex, municipality, month and easter.

As for mental health, there is a lockdown-dip followed by a recuperation for this outcome. However, compared to mental health consultations, the share of any primary care consultation displays a much more modest increase after the lockdown period for the two youngest age groups. For the age group 16-19, the total number of consultations falls throughout the school year 2020-202. Difference-in-difference estimates (Table 2) suggest that as of 2021, primary care consultations were unchanged for children aged 13-15 year and had fallen by 3.2 percent in the youngest age group, and 25.8 percent in the oldest age group. One explanation for the sharp fall in the oldest age group, is that prior to lockdown, sickness absence from high school (which this age group attends) beyond a low threshold had to be doctor certified. After lockdown, this requirement was removed, potentially changing the need for primary care services in this group quite substantially. As discussed in the main text, this is also likely to influence the results for mental health consultations. For the two youngest age groups, absence is certified by parents rather than doctors.

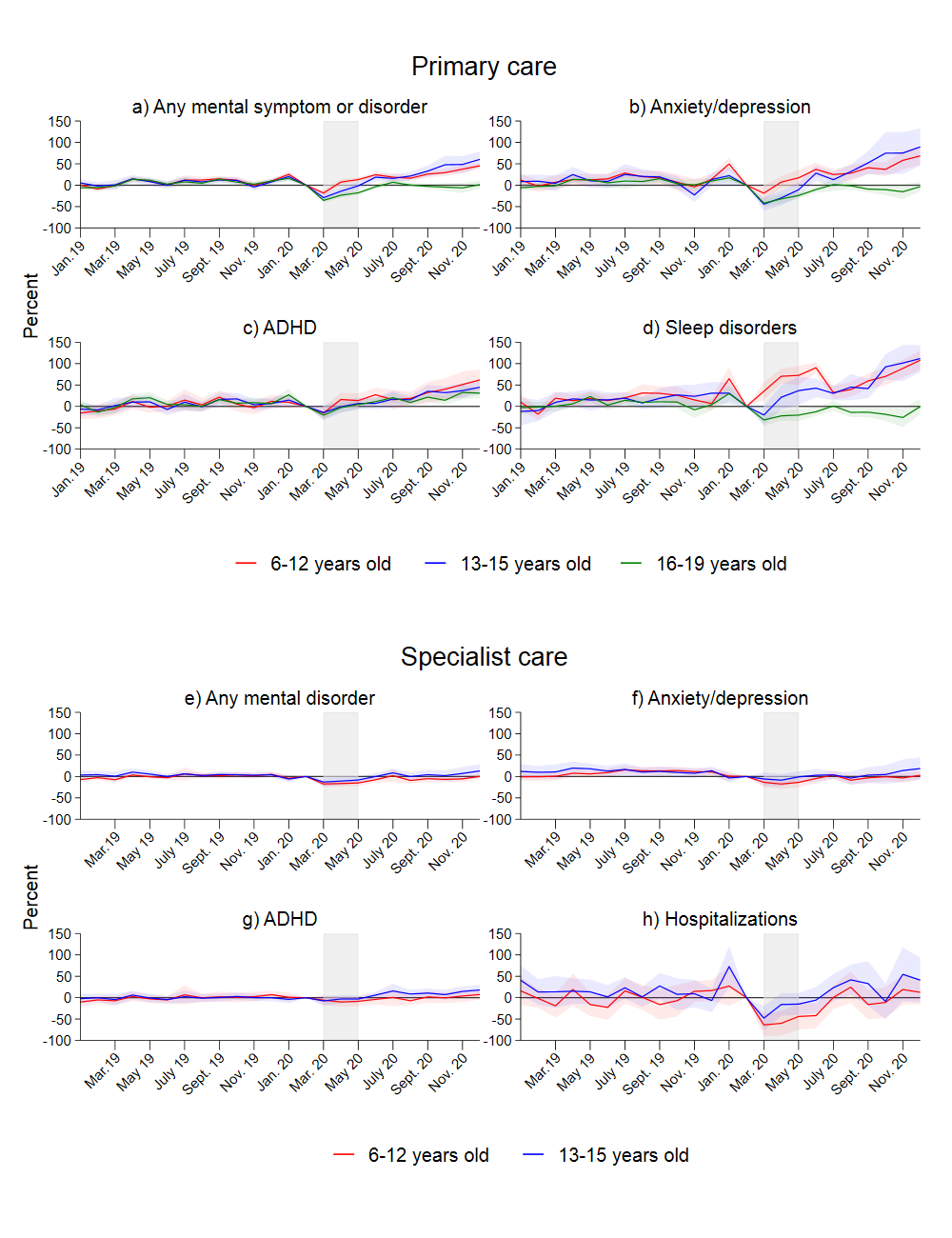

**Figure A.6:** Results from separate event study models for three age groups. Shortened observation window. Complete lines show coefficients, and shaded areas their 95% confidence intervals. Coefficients and confidence intervals are scaled to the pre-lockdown level in the main sample (see Table 1). The outcome is the monthly propensity to have at least one consultation of the type mentioned in the panel headers. Diagnoses are based on IPCD codes for primary care, and ICD-10 for specialist care (see Table A.1). The x-axis refers to the measurement time for the main sample. For the comparison sample, all measurements are taken 24 months earlier. Models control for duration in years.

1. For 2019, most person-month records will be included in both the control- and intervention cohort (albeit at different durations). We test whether the results are sensitive to this by reducing the observation period, so that no person month is included in both the main and comparison sample. The results are not sensitive to this (Figure A.6).
